# Using Large Language Models to Determine Reasons for Missed Colon Cancer Screening Follow-Up

**DOI:** 10.1101/2025.06.11.25329439

**Authors:** Christopher Y.K. Williams, Urmimala Sarkar, Julia Adler-Milstein, Lisa Rotenstein

## Abstract

**Importance:** Identifying reasons for missed preventive care, such as follow-up colonoscopy after an abnormal stool-based colon cancer screening test, is critical for quality improvement initiatives. However, manual chart review to extract this information from unstructured clinical notes is time-consuming and costly.

**Objective:** To determine whether a large language model (LLM) can accurately extract reasons for a lack of follow-up colonoscopy after abnormal outpatient fecal immunohistochemical test (FIT) or fecal occult blood test (FOBT).

**Design:** Cross-sectional study.

**Setting:** University of California, San Francisco (UCSF).

**Participants:** Adult patients aged 45 years or older with an abnormal outpatient FIT/FOBT between 2012 and 2024 who did not undergo a colonoscopy within 90 days of the abnormal test.

**Exposure:** We investigate the potential of an LLM to determine whether reasons for a lack of follow-up colonoscopy are documented in the clinical notes and whether an LLM can accurately classify those reasons into clinically meaningful categories.

**Main Outcomes and Measures:** Accuracy score was calculated to evaluate LLM performance against a 10% subsample manually classified by a physician reviewer.

**Results:** From a total of 2164 patients with abnormal FIT/FOBTs performed at UCSF during the study period, 355 (16.4%) underwent a colonoscopy within 90 days of the abnormal test. Among those who did not receive a colonoscopy within 90 days, 846 patients were eligible for the main analysis. Based on LLM categorization of patient note content, 270 (31.9%) patients did not have any reference to colonoscopy/colorectal cancer screening in their notes, 379 (44.8%) patients had mentions of colonoscopy/colorectal cancer screening without explicit reasons for not having a colonoscopy provided, and 197 (23.3%) patients had notes detailing explicit reasons for not having a colonoscopy. Overall LLM classification accuracy was 89.3%. The most common reasons for not having a colonoscopy included: Refused/not interested (n = 96; 35.2%), Comorbidities (n = 51; 18.7%), and Patient Unavailable (n = 46; 16.8%).

**Conclusions and Relevance:** This study suggests that an LLM can accurately identify and categorize reasons for the absence of follow-up colonoscopy after an abnormal FIT/FOBT. Our results suggest that LLMs have the potential to automate chart review for quality improvement initiatives.

## Introduction

Preventive care, which includes services such as cancer screening, vaccination, bone density measurement, and screening for common chronic diseases such as diabetes and hyperlipidemia,^1^ is an essential part of modern healthcare.^2^ In the United States, in response to specific preventive care measures recommended by the United States Preventive Services Task Force (USPSTF),^3^ delivery of preventive services has been incentivized through payments from payors. For example, in Medicare’s Merit-based Incentive Payment System (MIPS), better performance on specific quality measures is tied to financial payments.^4^ Similarly, value-based contracts incentivize quality measure performance across a variety of payor types, including commercial, Medicare, and Medicaid.^5^

There are two key challenges arising from this environment. First, the process of extracting data for - and reporting on - incentivized quality measures has created an administrative burden for clinicians and medical practices. For example, the cost to practices of reporting quality measures to MIPS is estimated at $12,811 per-physician.^6^ When manual abstraction of patient charts is required to report a quality metric, this amplifies the administrative burden and increases costs. Second, quality measurement necessitates further investigation into individual patient records in order to understand why they did not receive recommended care. Most of the data on missed preventive care reside in unstructured fields - free text - which makes it difficult to automate this investigative process. Following investigation, health systems must undertake targeted follow-up to close these care gaps. By identifying actionable opportunities to improve adherence to preventive care, teams of patient navigators, care coordinators and other personnel can be deployed to help facilitate care gap closure.^7^

With the recent emergence of large language models (LLMs), which are AI systems capable of processing and generating large quantities of human language, there is increasing interest in applying natural language processing-based approaches to both of these challenges.^8–17^ Previous studies have demonstrated success in using LLMs across various clinical tasks, including phenotyping patients, annotating pathology reports, clinical trial screening, identifying reasons for medication switching, and Emergency Department triage, among others.^13,14,16,18–21^ However, given the nascency of this field, existing literature evaluating the use of LLMs for quality improvement initiatives is limited.^22^ In particular, few prior studies have explored their ability to mine large amounts of healthcare data in ways that mimic the chart review processes that are central to many quality improvement efforts.^23^

To better understand how LLMs could be used to reduce administrative effort related to quality measure abstraction and identify reasons for lack of care gap closure, we undertook a pilot study in the context of a key preventive care recommendation: colorectal cancer (CRC) screening. As the second most common cause of cancer death in the United States, CRC is a major focus of preventive care.^24^ The USPSTF recommends that adults aged 45 to 75 be screened for CRC, with decisions for patients aged older than 75 made on an individual basis.^25^ Available non-invasive CRC screening tests include the guaiac-based fecal occult blood test (FOBT) and fecal immunochemical test (FIT), which detect microscopic amounts of blood in stool by targeting either heme (FOBT) or human globin (FIT).^26,27^ Offering these inexpensive tests as a screening option improves adherence to CRC screening and is associated with a 16% reduction in the relative risk of CRC mortality.^28,29^ Patients with an abnormal FOBT/FIT should have a follow-up colonoscopy performed, yet often this is delayed or missed, leading to a higher risk of later-stage CRC incidence and death.^30^ Identification of reasons for missed follow-up colonoscopy is therefore essential so that quality improvement interventions can be developed to act on reasons for missed follow up.^31,32^ However, extracting these reasons is often a costly and time-intensive process, requiring manual chart abstraction of relevant information from clinical notes, and consequently offers a prime opportunity to evaluate the use of LLMs for this task.^33–35^

In this study, after identifying patients who had not completed a colonoscopy within 90 days of an abnormal FIT/FOBT, we investigated the ability of an LLM to a) determine whether reasons for a lack of follow-up colonoscopy are documented in the clinical notes and b) accurately classify those reasons into clinically meaningful categories. These results speak to the ability of LLMs to serve as a substitute for manual chart review for quality improvement and could increase adherence to CRC screening by expediting team-based workflows to improve rates of colonoscopy completion after abnormal FIT/FOBT.

## Methods

### Data Source

The UCSF Information Commons contains deidentified structured clinical data as well as deidentified clinical text notes from all inpatient and ambulatory encounters at UCSF Health (a large academic medical center).^36^ The UCSF Institutional Review Board determined that use of this externally-certified, deidentified data within the UCSF Information Commons environment is not human subjects research and therefore this study was exempt from review.

### Identification of Study Cohort

In accordance with the USPSTF guidelines, we retrieved all adult patients aged 45 years or older from 2012 to 2024 with an abnormal (positive) Fecal Immunohistochemistry Test (FIT) or Fecal Occult Blood Test (FOBT) performed as an outpatient.^25^ We excluded FIT/FOBTs performed during an inpatient encounter, alongside FIT/FOBTs undertaken after the first abnormal (positive) test, because these FIT/FOBT tests were ordered for other reasons. We next retrieved the dates for subsequent colonoscopy procedures performed following an abnormal FIT/FOBT. We identified colonoscopies performed at UCSF using both procedure codes (CPT code: ‘45378’, ‘45379’, ‘45380’, ‘45381’, ‘45382’, ‘45388’, ‘45384’, ‘45385’, ‘45386’, ‘45389’, ‘45391’, ‘45392’, ‘45390’, ‘45393’, ‘45398’; HCPCS code: ‘G0105’, ‘G0121’) and review of colonoscopy-related procedures (Table S1). We calculated time from abnormal FIT/FOBT to colonoscopy and identified patients with no colonoscopy performed within 90 days of an abnormal FIT/FOBT. For these patients, we extracted de-identified clinical notes written within this 90-day period from the electronic health record. For each patient, we concatenated clinical notes, and patients with notes exceeding 31,500 tokens (corresponding to the 32,000 token context window of GPT-4-32K, with 500 tokens reserved for prompting and completion tokens) in combined length were excluded.

### LLM analysis

Through several iterations of prompt engineering, the LLM was prompted to classify each patient who had not had a colonoscopy within 90 days of an abnormal FIT/FOBT into one of the following three categories based on the text present in each patient’s clinical notes: 1) No relevant information available; 2) Relevant information available, but no explicit reasons for not having a colonoscopy: <free-text>; 3) Relevant information available and explicit reasons for not having a colonoscopy: <free-text>. To better characterize reasons for not having a colonoscopy, the free-text returned by the LLM for Categories 2 and 3 was further categorized into subcategories (Tables S2 & S3). Ten percent of LLM-generated labels and associated free-text were checked for accuracy by a physician reviewer.

## Results

We identified 2164 patients with abnormal FIT/FOBTs performed at UCSF Health in our study cohort, of which 355 (16.4%) underwent a colonoscopy within 90 days of the abnormal test. For the remaining 1809 patients who did not receive a colonoscopy within 90 days of an abnormal FIT/FOBT, 1062 (58.7%) patients had one or more clinical notes documented within 90 days following abnormal FIT/FOBT and 747 (41.2%) patients did not. A further 13 patients were identified with a colonoscopy report documented in their clinical notes and were excluded. Following concatenation of all notes for each patient and tokenization, 203 patients with notes exceeding 31,500 tokens were excluded. This left 846 patients with notes eligible for the main analysis (Table 1).

**Table 1.** Demographic characteristics of patients (n = 846) with no follow-up colonoscopy within 90 days of abnormal FIT/FOBT.

|  | Count (%) |
| --- | --- |
| **Sex** |  |
| Male | 439 (51.9) |
| Female | 407 (48.1) |
| **Race/Ethnicity** |  |
| White | 365 (43.1) |
| Asian | 224 (26.5) |
| Black or African American | 96 (11.3) |
| Latinx | 78 (9.2) |
| Multi-Race/Ethnicity | 28 (3.3) |
| Other | 27 (3.2) |
| Unknown/Declined | 11 (1.3) |
| Southwest Asian and North African | 8 (0.9) |
| Native Hawaiian or Other Pacific Islander | 7 (0.8) |
| Native American or Alaska Native | 2 (0.2) |
| **Age** |  |
| Median (IQR) |  |
| 64.8 years (56.3 – 72.1 years) |  |
| 45 to 55 years | 170 (20.1) |
| 55 to 65 years | 261 (30.9) |
| 65 to 75 years | 267 (31.6) |
| 75+ years | 148 (17.5) |

Using an LLM to classify patient notes, 270 (31.9%) patients did not have any reference to colonoscopy/colorectal cancer screening in their notes (Category 1), 379 (44.8%) patients had mentions of colonoscopy/colorectal cancer screening without explicit reasons for not having a colonoscopy provided (Category 2), and 197 (23.3%) patients had notes detailing explicit reasons for not having a colonoscopy (Category 3).

Following manual physician review of LLM classification accuracy, 88.9% (24/27) of patients in the manually reviewed Category 1 sample were confirmed to be correctly labelled. 89.2% (33/37) of patients in the Category 2 sample were confirmed as correct, with appropriate free-text extracted by the LLM in 100% (33/33) of cases. 90.0% (18/20) of patients in the Category 3 sample were confirmed as correct, with appropriate free-text extracted by the LLM in 94.4% (17/18) of cases. Combining the three samples, the overall LLM classification accuracy was 89.3%.

### Subcategory classification

Among the 379 patients with mentions of colonoscopy/colorectal cancer screening without explicit reasons for not having a colonoscopy provided (Category 2), 183 (48.3%) were referred for a colonoscopy, but there was no documented evidence in their notes of the colonoscopy being scheduled, while 101 (26.6%) had documented evidence of their colonoscopy being scheduled (Table 2). The most common reasons provided for the 197 patients with explicit reasons for not having a colonoscopy (Category 3) included: Refused/not interested (n = 96; 35.2%), Comorbidities (n = 51; 18.7%), Patient Unavailable (n = 46; 16.8%), Other (n = 24; 8.8%), and Mental Health/Psychiatric reason (n = 13; 4.8%) (Table 3).

**Table 2.** Classification of 379 patients with mentions of colonoscopy/colorectal cancer screening without explicit reasons for not having a follow-up colonoscopy provided.

| **Subcategory** | **Count (%)** |
| --- | --- |
| 'Positive Fecal Immunochemical Test (FIT) mentioned/identified and patient referred to Gastroenterology/colonoscopy ordered, but colonoscopy not scheduled' | 183 (48.3) |
| 'Fecal Immunochemical Test (FIT) mentioned/identified and patient scheduled for colonoscopy' | 101 (26.6) |
| 'Other' | 72 (19.0) |
| 'Positive Fecal Immunochemical Test (FIT) mentioned/identified and patient informed but no referral to Gastroenterology or ordering for colonoscopy' | 16 (4.2) |
| 'Fecal Immunochemical Test (FIT) repeated instead of requesting/scheduling colonoscopy' | 7 (1.85) |
| 'Positive Fecal Immunochemical Test (FIT) mentioned/identified but patient not informed and no referral to Gastroenterology or ordering of colonoscopy' | 0 (0) |
| **Total** | **379** |

**Table 3.** Classification of reasons extracted by the LLM for the 197 patients with explicit reasons for not having a colonoscopy. Each patient could be assigned one or more subcategories (e.g. if multiple reasons are provided in their notes).

| **Subcategory** | **Definition** | **Count (%)** |
| --- | --- | --- |
| Refused/not interested | A provider or staff member documented that the patient refused or is not interested in colonoscopy | 96 (35.2) |
| Comorbidities | The chart states that the patient has comorbid health conditions which make them unfit for colonoscopy/colonoscopy is contraindicated - for example, life expectancy of less than 10 years, Stage 4 cancer, dementia | 51 (18.7) |
| Patient unavailable | A provider or staff member documented that the patient does not have availability for colonoscopy due to scheduling issues - for example, upcoming travel or work/family obligations. | 46 (16.8) |
| Other |  | 24 (8.79) |
| Mental health/Psychiatric reason | A provider or staff member documented that the patient has mental health issues that would interfere with colonoscopy. Use this label only if relevant to colorectal cancer screening continuum | 13 (4.76) |
| Diet and laxative instructions not followed | A provider or staff member documented that the patient did not follow diet and laxative instructions - for example, the patient did not go on clear-liquid diet or did not drink laxative as prescribed | 10 (3.66) |
| Low health literacy (patient does not understand) | A provider or staff member documented that the patient could not verbalize understanding of colonoscopy and/or the positive FIT result. Use this label if the health care team attempted to educate patient on colonoscopy and/or positive FIT but were unable to reach an understanding. | 9 (3.30) |
| Transportation | A provider or staff member documented that the patient is unable to travel to the hospital for colonoscopy due to lack of transportation | 8 (2.93) |
| Social isolation | A provider or staff member documented that the patient does not have adequate social support to assist with post-colonoscopy recovery- for example, the patient needs assistance at home and no house member is able to render care. Use this label only if relevant to colorectal cancer screening continuum. | 4 (1.47) |
| Unable to reach | Clinic staff is unable to contact patient. Also use this label if the clinic staff has documented that the patient's phone number and emergency contact numbers do not reach the patient. | 4 (1.47) |
| Not in health network | The chart states that the patient is no longer being seen at our health network - for example, wording like "transferring care to Kaiser". | 4 (1.47) |
| Prescription laxative issue | A provider or staff member documented that the patient did not pick up their prescription laxative - for example, the patient was unaware of prescription, unsure of which pharmacy to go to, or the pharmacy they went to did not have their prescription ready. | 2 (0.73) |
| Substance use | A provider or staff member documented that the patient has substance use issues that would interfere with colonoscopy. Use this label only if relevant to colorectal cancer screening continuum | 1 (0.37) |
| Homelessness | The chart states that the patient is experiencing homelessness. | 1 (0.37) |

Among the 10% sample of patients in Category 2 and 3 manually reviewed by a physician to validate appropriate subcategory classification by the LLM, 25/37 (67.6%) of Category 2 subcategory labels were accurate, while 16 of 20 (80.0%) of Category 3 subcategory labels manually reviewed by a physician were accurate. Comparisons of physician- and LLM-classified subcategory labels are displayed in Supplementary Tables S4 & S5.

## Discussion

In this study of 846 patients without follow-up colonoscopy within 90 days of an abnormal FIT/FOBT, we demonstrate that an LLM can both identify whether reasons for a lack of follow-up colonoscopy are documented in the clinical notes and accurately classify those reasons into clinically meaningful categories. Only 23.4% of patients had explicit reasons for not completing a follow-up colonoscopy described in their notes; 31.9% of patients did not have any reference to colonoscopy/CRC screening recorded in their notes, while 44.8% had mentions of colonoscopy/CRC screening but no explicit reasons documented for not having a colonoscopy.

LLM classification achieved an overall accuracy of 89.3% on physician review, highlighting the potential utility of LLMs to inform quality improvement efforts.

Among those patients who did have explicit reasons for their lack of follow-up colonoscopy documented, the most frequently identified reasons were the patient not consenting to the procedure (35.2%) and contraindication to colonoscopy due to patient comorbidities (18.7%). Among other, potentially addressable, reasons for not having a colonoscopy, social determinants of health played a prominent role, with patient availability (16.8%), low health literacy (3.3%), transportation (2.9%), and social isolation (1.5%) reported as common reasons. Similar findings have been reported at several other institutions.^33–35,37^ In a retrospective study of 12 Veterans Affairs sites in southern California, 35.2% of patients declined colonoscopy following abnormal FIT, while 12.1% had scheduling issues and 11.2% did not show for their colonoscopy appointment.^37^ Meanwhile, in a mixed-methods study conducted at a large safety-net healthcare system, the most frequently documented reasons included declined colonoscopies, competing health issues, lack of transportation and patient no-show to colonoscopy.^33^ These combined findings suggest that, although a lack of follow-up colonoscopy may be appropriate for certain patients, enhancing availability and access to colonoscopy services is essential to ensure patients can obtain the procedure where indicated.

The use of large language models in healthcare is a nascent, but rapidly evolving, field. Early studies showing strong performance of LLMs when answering clinical questions in curated text such as the United States Medical Licensing Exam have more recently been followed by studies evaluating this performance on real-world clinical notes.^10,15,19^ The potential benefits of this technology for quality improvement, including the extraction of patient information for quality measurement and reporting, are beginning to be realized.^38^ One recent study showed that a natural language processing-based model achieved good performance when tasked with determining the NIH Stroke Scale score of adult patients presenting with acute stroke.^22^ Another found that an automated machine learning model could accurately apply the Mayo endoscopic subscore to colonoscopy reports to classify disease activity in ulcerative colitis.^39^ To our knowledge, our study is the first to evaluate the use of LLMs to identify why gaps in patient care exist. This is an important first step in understanding the potential future uses of LLMs in quality improvement. Our findings suggest that LLMs may significantly streamline information extraction from the electronic health record, thereby reducing the cost of manual chart abstraction for quality measures and providing addressable insights that can be targeted to improve population health.^38^

There are several limitations to this study. First, the data used in this study was limited to the deidentified clinical notes available for each patient. Further information on reasons for not having a follow-up colonoscopy may be found in patient in-basket messages to their providers and future studies should seek to evaluate this additional source of data. Second, we only examined the 90-day period following abnormal FIT/FOBT; patients may have received colonoscopies after this period. Third, due to the limited context-window of the LLM, we limited our cohort to patients with clinical notes under 31,500 tokens in total length, leading to the exclusion of a minority (n = 203) of patients. These patients may have more complex health requirements and, consequently, it is important to consider alternative approaches when applying these models to larger corpuses of text for more complex patients. Here, we concatenated all notes into a single corpus for input into the LLM; although a note-by-note classification would overcome the context window limitations for most patients, it is unclear whether such an approach would have similar accuracy given the consequent lack of context (from other notes) provided when each note is evaluated in isolation. Third, only one LLM (GPT-4-32K) was evaluated and model performance may improve with more comprehensive prompt engineering.^40^ Fourth, this was a single-center study focusing on only one type of quality measure related to abnormal stool test follow-up; future studies should investigate the ability of LLMs to conduct chart review and summarization at scale across different institutions and for other types of screening guidelines.

### Conclusion

In this study of 846 patients who did not have a follow-up colonoscopy within 90 days of an abnormal FIT/FOBT, we found that an LLM can both identify whether reasons for a lack of follow-up colonoscopy are documented in patients’ clinical notes and accurately classify those reasons into clinically meaningful categories. These results suggest the potential ability of LLMs to facilitate chart review and inform quality improvement efforts at scale. Future studies should investigate the application of LLMs to further characterize gaps in care and the reasons behind those gaps.

## Data Availability

All data supporting the findings described in this manuscript are available in the article, Supplementary Information, or from the corresponding author upon request. The UCSF Information Commons database is available to individuals affiliated with UCSF who can contact the UCSF’s Clinical and Translational Science Institute (CTSI) or the UCSF’s Information Commons team for more information.

## Acknowledgements

This study was funded by The Doctors Company. The funder had no role in the study design, data collection, analysis, or interpretation of results.

The authors acknowledge the use of the UCSF Information Commons computational research platform, developed and supported by UCSF Bakar Computational Health Sciences Institute. The authors also thank the UCSF AI Tiger Team, Academic Research Services, Research Information Technology, and the Chancellor’s Task Force for Generative AI for their software development, analytical and technical support related to the use of Versa API gateway (the UCSF secure implementation of large language models and generative AI via API gateway), Versa chat (the chat user interface), and related data asset and services.

Dr Christopher Y.K. Williams had full access to all the data in the study and takes responsibility for the integrity of the data and the accuracy of the data analysis.

