## Supplemental File 1 for "Using Large Language Models to Determine Reasons for Missed Colon Cancer Screening Follow-Up"

**GPT-4-32K prompt**

Main label (Category 1, 2, or 3) categorization:

*"Below are a series of notes of a patient with an abnormal Fecal Occult Blood Test (FOBT)/Fecal Immunohistochemistry Test (FIT) who should have received a colonoscopy but did not. Each note is separated by the following delimiter: ---///---*

*###START###*

*<NOTE_TEXT>*

*###END##*

*Using the above series of notes, identify any mentions of colonoscopy or other investigations screening for bowel/colorectal cancer and also provide reasons (if any) for the patient not having a colonoscopy despite having an abnormal FOBT/FIT test. Return one of the following: {\'0: No relevant information available', \'1: Relevant information available, but NO explicit reasons for not having a colonoscopy: <TEXT>', \'2: Relevant information available AND explicit reasons for not having a colonoscopy: <REASONS>'}"*

Category 2: Sublabel categorization:

*“Below are reasons extracted from the electronic health record for why a patient who had an abnormal Fecal Occult BLood Test (FOBT)/Fecal Immunohistochemistry Test (FIT), who should have received a colonoscopy, did not have a colonoscopy.*

*Reasons:*

*<TEXT>*

*Categorize the reasons using one label from the following schema: '*

*0: 'Positive Fecal Immunochemical Test (FIT) mentioned/identified but patient not informed and no referral to Gastroenterology or ordering of colonoscopy',*

*1: 'Positive Fecal Immunochemical Test (FIT) mentioned/identified and patient informed but no referral to Gastroenterology or ordering for colonoscopy',*

*2: 'Positive Fecal Immunochemical Test (FIT) mentioned/identified and patient referred to Gastroenterology/colonoscopy ordered, but colonoscopy not scheduled',*

*3: 'Fecal Immunochemical Test (FIT) mentioned/identified and patient scheduled for colonoscopy',*

*4: 'Fecal Immunochemical Test (FIT) repeated instead of requesting/scheduling colonoscopy',*

*5: 'Other (give reasons)'*

*Return only one label.”*

Category 2: Sublabel categorization:

*“Below are reasons extracted from the electronic health record for why a patient who had an abnormal Fecal Occult BLood Test (FOBT)/Fecal Immunohistochemistry Test (FIT), who should have received a colonoscopy, did not have a colonoscopy.*

*Reasons:*

*<TEXT>*

*Categorize the reasons using labels (one or more) from the following schema: '*

*0: 'Unable to reach: Clinic staff is unable to contact patient. Also use this label if the clinic staff has documented that the patient’s phone number and emergency contact numbers do not reach the patient.',*

*1: 'Left voice message: Clinic staff documented that they left voicemail for patient at one of the numbers in the chart.',*

*2: 'Refused, not interested: A provider or staff member documented that the patient refused or is not interested in colonoscopy.',*

*3: 'Not in health network: The chart states that the patient is no longer being seen at our health network - for example, wording like “transferring care to Kaiser”.',*

*4: 'Comorbidities: The chart states that the patient has comorbid health conditions which make them unfit for colonoscopy/colonoscopy is contraindicated - for example, life expectancy of less than 10 years, Stage 4 cancer, dementia.',*

*5: 'Transportation: A provider or staff member documented that the patient is unable to travel to the hospital for colonoscopy due to lack of transportation.',*

*6: 'Unavailable (family, work etc): A provider or staff member documented that the patient does not have availability for colonoscopy due to scheduling issues - for example, upcoming travel or work/family obligations.',*

*7: 'Social isolation: A provider or staff member documented that the patient does not have adequate social support to assist with post-colonoscopy recovery- for example, the patient needs assistance at home and no house member is able to render care. Use this label only if relevant to colorectal cancer screening continuum.',*

*8: 'Mental health, psych: A provider or staff member documented that the patient has mental health issues that would interfere with colonoscopy. Use this label only if relevant to colorectal cancer screening continuum.',*

*9: 'Substance use : A provider or staff member documented that the patient has substance use issues that would interfere with colonoscopy. Use this label only if relevant to colorectal cancer screening continuum.',*

*10: 'Homelessness: The chart states that the patient is experiencing homelessness.',*

*11: 'Low health literacy (patient does not understand): A provider or staff member documented that the patient could not verbalize understanding of colonoscopy and/or the positive FIT result. Use this label if the health care team attempted to educate patient on colonoscopy and/or positive FIT but were unable to reach an understanding.',*

*12: 'Prescription laxative issue: A provider or staff member documented that the patient did not pick up their prescription laxative - for example, the patient was unaware of prescription, unsure of which pharmacy to go to, or the pharmacy they went to did not have their prescription ready.',*

*13: 'Diet and laxative instructions not followed: A provider or staff member documented that the patient did not follow diet and laxative instructions - for example, the patient did not go on clear-liquid diet or did not drink laxative as prescribed.',*

*14: 'Other'*

*Return the output in the form of a python list of labels.”*

**Included colonoscopy-related procedures**

'PR COLONOSCOPY STOMA W/ENDOSCOPIC MUCOSAL RESCJ'

'INACTIVE PEDIATRICS COLONOSCOPY WITH OR WITHOUT BIOPSY'

'PR COLONOSCOPY W/BIOPSY SINGLE/MULTIPLE'

'ENDO ADULT COLONOSCOPY WITH BIOPSY'

'ENDO ADULT COLONOSCOPY DIAGNOSTIC'

'ENDO ADULT COLONOSCOPY WITH INJECTION THERAPY'

'ENDO PEDS EGD AND COLONOSCOPY BIOPSY; DISACCHARIDASE ANALYSIS'

'ENDO PEDS COLONOSCOPY WITH COAGULATION'

'ENDO ADULT EGD/COLONOSCOPY'

'ENDOSCOPY, COLON, DIAGNOSTIC'

'ENDO ADULT COLONOSCOPY WITH POLYPECTOMY'

'COLONOSCOPY'

'ENDO PEDS COLONOSCOPY DIAGNOSTIC'

'PR COLONOSCOPY FLX DX W/COLLJ SPEC WHEN PFRMD'

'ENDO ADULT COLONOSCOPY WITH COAGULATION'

'COLONOSCOPY W/ BIOPSIES'

'PR COLONOSCOPY FLX W/ENDOSCOPIC MUCOSAL RESECTION'

'PR COLONOSCOPY FLX ABLATION TUMOR POLYP/OTHER LES'

'PR COLONOSCOPY STOMA W/BIOPSY SINGLE/MULTIPLE'

'COLONOSCOPY W/ POLYPECTOMY'

'ENDO ADULT COLONOSCOPY WITH ENDOSCOPIC MUCOSAL RESECTION'

'PEDIATRIC COLONOSCOPY'

'PEDIATRICS ESOPHAGOGASTRODUODENOSCOPY (EGD) AND COLONOSCOPY'

'ENDO ADULT COLONOSCOPY WITH FECAL MICROBIOTA INFUSION'

'ENDO PEDS COLONOSCOPY WITH POLYPECTOMY'

'ENDO ADULT COLONOSCOPY WITH ULTRASOUND'

'ENDO PEDS COLONOSCOPY WITH BIOPSY'

'PR COLON CA SCRN NOT HI RSK IND'

'COLONOSCOPY POLYPECTOMY WITH CAUTERIZATION'

**Table S1.** Colonoscopy procedures included following manual review.

**Subcategory label**

'Positive Fecal Immunochemical Test (FIT) mentioned/identified but patient not informed and no referral to Gastroenterology or ordering of colonoscopy'

'Positive Fecal Immunochemical Test (FIT) mentioned/identified and patient informed but no referral to Gastroenterology or ordering for colonoscopy'

'Positive Fecal Immunochemical Test (FIT) mentioned/identified and patient referred to Gastroenterology/colonoscopy ordered, but colonoscopy not scheduled'

'Fecal Immunochemical Test (FIT) mentioned/identified and patient scheduled for colonoscopy'

'Fecal Immunochemical Test (FIT) repeated instead of requesting/scheduling colonoscopy'

‘Other’

**Table S2**. List of subcategory labels used to classify patients whose notes mentioned colonoscopy/colorectal cancer screening but did not contain explicit reasons for not having a follow-up colonoscopy after abnormal FIT/FOBT.

| **Subcategory label** | **Description** |
| --- | --- |
| Unable to reach | *Clinic staff is unable to contact patient. Also use this label if the clinic staff has documented that the patient’s phone number and emergency contact numbers do not reach the patient.,* |
| Left voice message | *Clinic staff documented that they left voicemail for patient at one of the numbers in the chart* |
| Refused, not interested | *A provider or staff member documented that the patient refused or is not interested in colonoscopy* |
| Not in health network | *The chart states that the patient is no longer being seen at our health network - for example, wording like “transferring care to Kaiser”.* |
| Comorbidities | *The chart states that the patient has comorbid health conditions which make them unfit for colonoscopy/colonoscopy is contraindicated - for example, life expectancy of less than 10 years, Stage 4 cancer, dementia.'* |
| Transportation | *A provider or staff member documented that the patient is unable to travel to the hospital for colonoscopy due to lack of transportation.'* |
| Unavailable (family, work etc.) | *A provider or staff member documented that the patient does not have availability for colonoscopy due to scheduling issues - for example, upcoming travel or work/family obligations.'* |
| Social isolation | *A provider or staff member documented that the patient does not have adequate social support to assist with post-colonoscopy recovery- for example, the patient needs assistance at home and no house member is able to render care. Use this label only if relevant to colorectal cancer screening continuum.* |
| Mental health, psych | *A provider or staff member documented that the patient has mental health issues that would interfere with colonoscopy. Use this label only if relevant to colorectal cancer screening continuum.* |
| Substance use | *A provider or staff member documented that the patient has substance use issues that would interfere with colonoscopy. Use this label only if relevant to colorectal cancer screening continuum* |
| Homelessness | *The chart states that the patient is experiencing homelessness* |
| Low health literacy (patient does not understand) | *A provider or staff member documented that the patient could not verbalize understanding of colonoscopy and/or the positive FIT result. Use this label if the health care team attempted to educate patient on colonoscopy and/or positive FIT but were unable to reach an understanding* |
| Prescription laxative issue | *A provider or staff member documented that the patient did not pick up their prescription laxative - for example, the patient was unaware of prescription, unsure of which pharmacy to go to, or the pharmacy they went to did not have their prescription ready.'* |
| Diet and laxative instructions not followed | *A provider or staff member documented that the patient did not follow diet and laxative instructions - for example, the patient did not go on clear-liquid diet or did not drink laxative as prescribed* |
| Other | *n/a* |

**Table S3**. List of subcategory labels used to classify extracted reasons for missed follow-up colonoscopy after abnormal FIT/FOBT.

| **Physician-assigned subcategory label (ground-truth)** | **LLM-assigned subcategory label** | **Count** |
| --- | --- | --- |
| 1: ‘Positive Fecal Immunochemical Test (FIT) mentioned/identified and patient informed but no referral to Gastroenterology or ordering for colonoscopy’ | 2: 'Positive Fecal Immunochemical Test (FIT) mentioned/identified and patient referred to Gastroenterology/colonoscopy ordered, but colonoscopy not scheduled' | 4 |
| 5: ‘Other’ | 1: ‘Positive Fecal Immunochemical Test (FIT) mentioned/identified and patient informed but no referral to Gastroenterology or ordering for colonoscopy’ | 2 |
| 1: ‘Positive Fecal Immunochemical Test (FIT) mentioned/identified and patient informed but no referral to Gastroenterology or ordering for colonoscopy’ | 5: ‘Other’ | 2 |
| 4: ‘Fecal Immunochemical Test (FIT) repeated instead of requesting/scheduling colonoscopy’ | 3: ‘Fecal Immunochemical Test (FIT) mentioned/identified and patient scheduled for colonoscopy’ | 1 |
| 3: ‘Fecal Immunochemical Test (FIT) mentioned/identified and patient scheduled for colonoscopy’ | 2: ‘Positive Fecal Immunochemical Test (FIT) mentioned/identified and patient referred to Gastroenterology/colonoscopy ordered, but colonoscopy not scheduled’ | 1 |
| 2: ‘Positive Fecal Immunochemical Test (FIT) mentioned/identified and patient referred to Gastroenterology/colonoscopy ordered, but colonoscopy not scheduled’ | 4: ‘Fecal Immunochemical Test (FIT) repeated instead of requesting/scheduling colonoscopy’ | 1 |
| 4: ‘Fecal Immunochemical Test (FIT) repeated instead of requesting/scheduling colonoscopy’ | 5: ‘Other’ | 1 |

**Table S4**. Breakdown of subcategory labels misclassified by the LLM in Category 2, compared to the ground-truth label assigned by physician reviewer.

| **Physician-assigned subcategory label(s) (ground-truth)** | **LLM-assigned subcategory label(s)** | **Count** |
| --- | --- | --- |
| 6: ‘Unavailable (family, work etc): A provider or staff member documented that the patient does not have availability for colonoscopy due to scheduling issues - for example, upcoming travel or work/family obligations.’ | 14: ‘Other’ | 2 |
| 2: 'Refused, not interested: A provider or staff member documented that the patient refused or is not interested in colonoscopy.'  AND  6: ‘Unavailable (family, work etc): A provider or staff member documented that the patient does not have availability for colonoscopy due to scheduling issues - for example, upcoming travel or work/family obligations.’} | 6: ‘Unavailable (family, work etc): A provider or staff member documented that the patient does not have availability for colonoscopy due to scheduling issues - for example, upcoming travel or work/family obligations.’ | 1 |
| 2: 'Refused, not interested: A provider or staff member documented that the patient refused or is not interested in colonoscopy.'  AND  4: ‘Comorbidities: The chart states that the patient has comorbid health conditions which make them unfit for colonoscopy/colonoscopy is contraindicated - for example, life expectancy of less than 10 years, Stage 4 cancer, dementia.’ AND  6: ‘Unavailable (family, work etc): A provider or staff member documented that the patient does not have availability for colonoscopy due to scheduling issues - for example, upcoming travel or work/family obligations.’} | 4: ‘Comorbidities: The chart states that the patient has comorbid health conditions which make them unfit for colonoscopy/colonoscopy is contraindicated - for example, life expectancy of less than 10 years, Stage 4 cancer, dementia.’ | 1 |

**Table S5**. Breakdown of subcategory labels misclassified by the LLM in Category 3, compared to the ground-truth label assigned by physician reviewer.
