## Supplementary material for "Using Large Language Models to Determine Reasons for Missed Colon Cancer Screening Follow-Up": Figure 1

- 747 patients with no available clinical notes
- 13 patients with colonoscopy report documented in clinical notes
- 203 patients with notes exceeding 31,500 tokens in length
- 355 (16.4%) underwent colonoscopy within 90 days of abnormal FOBT/FIT

846 patients eligible for main analysis

- 1100 abnormal FOBT/FIT from patients under 45 years
- 207 repeat tests

1809 (83.6%) without colonoscopy within 90 days of abnormal FOBT/FIT

2164 abnormal FOBT/FIT

3471 abnormal FOBT/FIT in total

50,163 total outpatient FOBT/FIT performed at UCSF between 2012-2023

Exclude

Exclude

Exclude

197 patients with notes detailing explicit reasons for not having a colonoscopy (Category 3)

379 patients with mentions of colonoscopy/colorectal cancer screening without explicit reasons for not having a colonoscopy provided (Category 2)

270 patients with no reference to colonoscopy/colorectal cancer screening in their notes (Category 1)

**Figure 1.** Flowchart of included patients
